# TO DETERMINE THE CLIENT RELATED FACTORS THAT INFLUENCE UPTAKE OF MATERNITY SERVICES BY WOMEN OF CHILDBEARING AGE ATTENDING LEVEL THREE HEALTH FACILITIES IN IMENTI SOUTH, MERU COUNTY

**DOI:** 10.1101/2024.06.20.24309021

**Authors:** Fridah Kawira Muchunku, Jane Karonjo, David Muya

## Abstract

The well-being of women before, during, and after delivery is referred to as “maternal health”. Numerous lives may be saved by maternal health care services despite the fact that millions of women of reproductive age die during pregnancy, delivery, and the postpartum period. More women are able to prevent short and long-term maternal disabilities and illnesses such as fistulas, infertility and depression. Kenya has developed a number of policies to improve maternal health. One such policy is the free maternity services policy that allows all pregnant mothers to have access to free maternity services in government facilities across the country. Free maternity services policy allows for provision of free delivery services to expectant women in health facilities run by the Kenyan Government. 189 nations ratified the Millennium Declaration and pledged to accomplish eight objectives by the turn of the twenty-first century. The study was a cross-sectional descriptive research design targeting population of 51,770 women of childbearing age in Imenti South Sub County. The sample size consisted of 100 respondents (women of childbearing aged between 18-49 years). The study employed qualitative and quantitative sampling methods. The sampling procedure was simple random sampling and purposive sampling. Data collection instruments were by questionnaires and observation schedule. Data was analyzed by use of descriptive statistics and results displayed using tables, bar charts, pie charts and presented using percentages, frequencies, means and standard deviations. Analysis was done using SPSS version 25. Uptake of maternity services. The negative attitude of health care workers greatly influenced Uptake of maternity services. Limited uptake was also linked to poor development of health infrastructure and usage of the facility, which ultimately influenced Uptake of maternity services. Awareness did not match Uptake of maternity services. The study concluded that social demographic factors, client related factors and health facility-related factors can influence the success or failure of Uptake of maternity services by women. It recommended a rights-based approach when designing and delivering free maternity services, strengthening client satisfaction programs and adopting a broader bottom-up approach when engaging rural women to ensure effective development of health infrastructure and usage of the facility. further suggestions for future research was recommended, including studies that explore attitudes and behaviours in varied settings, factors promoting positive attitudes and behaviours, and the effectiveness of interventions to address negative patient experiences in both three-level and other health facilities in Kenya.

## 1.0 Introduction

### 1.1 Background of the Study

The well-being of women before, during, and after delivery is referred to as “maternal health” (Ronsman & Graham, 2006). According to the World Health Organization [WHO] (2016) a significant number of women die due to difficulties during pregnancy and delivery, many of which are curable or avoidable. Nearly 75 percent of all maternal fatalities are caused by infections, excessive bleeding, complications from botched abortion and delivery, as well as pre-eclampsia and eclampsia.

Kenya has made enormous efforts towards achieving the SDG number three, that is improving maternal health. Among the measures in place to improve maternal health in Kenya is the free maternity services policy initiative adopted in 2013, according to USAID (2013) in agreement with the policy, all government owned health facilities were to offer free delivery services. For every delivery that occurred in a level two and three health facility, the Government would reimburse Ksh. 2,500 though the Health Sector Services Fund [HSSF]. In level four, five and six the government would reimburse Ksh. 5,000 through the Hospital Management Services Fund [HMSF].

The World Health Organization (2015) report states that globally, uptake of maternity services has improved from 62 percent to almost 80 percent between the years 2000 -2017 though the rate of progress varies across regions. The report further showed that Central and south Asia were the most improved regions with 77 percent of mothers accessing maternity services. According to WHO (2018) report, worldwide around 20 percent of the population are able to afford the care of a doctor or a midwife during pregnancy, delivery and postpartum period. The report further indicates that in under developed nations less than 40 percent of the populations are able to afford these appointments.

### 1.2 Statement of the Problem

Globally, about 80 percent of pregnant women have access to maternity services, according to a WHO (2015) report. The report further states that uptake of maternity services is a step towards achievement of the SDG specifically improving maternal health. In addition, more women are able to prevent short and long-term maternal disabilities and illnesses such as fistulas, infertility and depression. According to WHO (2015) access to maternity services in Sub Saharan Africa remains a challenge. Only one fifth of pregnant women have access to maternity services. In addition, KHDS (2014) report indicates that in Kenya only 62 percent of expectant mothers seek maternal care during the course of their pregnancy. Majority of these deliveries occur in level four and level five health facilities and in private health facilities. Less than twenty percent (20%) of these deliveries occurs in level three and level two health facilities.

### 1.3 Justification of the Study

According to a research published by WHO (2018) the use of maternity care is a step toward the accomplishment of the Sustainable Development Goals. The study goes on to say that a specific emphasis has been placed on Sub-Saharan Africa because it is the area with the lowest uptake of maternity services and the highest rate of maternal fatalities. Therefore, each country has established a unique set of strategies for enhancing maternal health care.

When it comes to addressing the issue of removing or subsidizing charges spent as fees at government health facilities in order to increase the use of maternity care in Kenya, the government has developed a planned policy. In a 2014 study, the Kenya Demographic and Health Survey [KDHS] said that the maternal death rate should be reduced to levels that are considered globally acceptable. As a result, the government committed to putting the Free Maternity Services policy into effect in order to reduce maternal mortality.

## 2.0 Literature Review

### 2.1 Client related factors: quality of care, healthcare worker’s attitude, ignorance

(Gathigah, 2013). Asserts that, despite the provision of free maternity care, the use of government health facilities in Kenya continues to be insufficiently increased. Women continue to give birth in private health facilities or seek services of traditional birth attendants. Gathigah further argues that according to a survey on utilization of government health facilities, fifty six of those interviewed said that they avoid going to government facilities because they are congested and the quality of service is poor. Fifteen per cent said that they avoided government facilities because of the negative attitudes and mistreatment from health care providers, while another twenty nine per cent mentioned a range of issues, including physical distance, cultural effect, and ignorance, as reasons for not utilizing government health facilities.

According to (Harding, 2010). Approximately 32 percent of pregnant women in Sub-Saharan Africa give birth in public health facilities, with the remaining 68 percent giving birth in private health facilities, which include home deliveries. (Harding, 2010). further argues when comparing private health facilities to public hospitals, private health facilities provide greater privacy, respect, and value for money. Issues such as abuse by health-care staff and bad treatments have harmed the reputation of public hospitals.

As with any other health-care delivery system, the utilization of maternal care in Africa is primarily dependent on the health-care administrators ensuring that it is equally dispersed across the country, particularly in rural places. According to the authors (Koblinsky & Campbell, 2003). There is a huge disparity between the provisions of prenatal and postnatal care in rural and urban areas in Egypt, for example. Obstetricians, physicians, and hospitals are few in Egypt’s rural, mostly agricultural areas, where the population is concentrated. As a result, the few facilities and resources that are available become congested, and some women choose not to take advantage of them, resulting in their inaccessibility owing to the high population density seen in urban regions. In contrast, since 2005, investments in maternity care in Egypt’s metropolitan districts have expanded dramatically, signaling a significant shift away from the country’s rural parts. The number of women seeking maternity care at public hospitals in metropolitan regions has increased considerably in recent years, owing to increased investment in the number of hospitals and the number of healthcare staff in the area.

Long before the advent of the modern era, Kenya was afflicted by high rates of maternal illness and mortality. Mothers die at a rate of 488 per 100,000 live births, according to the latest current estimates. This is much higher than the Millennium Development Goal of 147 deaths per 100,000 live births by 2015.

Approximately 80% of these births were administered by unqualified individuals who were ill-equipped to deal with complications that arose during pregnancy and delivery. There was also a general shortage of infrastructure to manage deliveries, which lies at the heart of the problem (Kenya National Commission on Human Rights, 2012). Aiming to increase the number of hospital births while concurrently lowering maternal mortality, Kenya implemented free maternal health care for all women in 2013. A number of obstacles stand in the way of this project, including insufficient funding and investment in new infrastructure, a paucity of equipment, and a lack of qualified employees to carry it out effectively and efficiently (Kenya National Commission for Human Rights, 2013).

In the opinion of (Bourbonnais, 2013). Women avoid using public health facilities during pregnancy and delivery because they have experienced abuse, mistreatment, and neglect on the part of health care providers, as well as poor service quality. A further point of contention is that government health facilities are not sympathetic to the cultural peculiarities of its patients, according to (Gathigah. 2013). While the public system does a good job of accommodating cultural and religious beliefs, it falls short in addressing the requirements of its citizens.

Approximately 51.9 percent of Kenyan women had access to competent delivery services, according to a study performed by (Chuma & Maina, 2013). Hospitals provide the bulk of these births, with primary health care facilities accounting for less than 9 percent of all births. There is anecdotal evidence, according to the authors, that primary health-care institutions are underused because they are seen to deliver low-quality services, have inadequate infrastructure, do not provide emergency services, and are ignored by the local population.

## 3.0 Research Methodology

The study used cross-sectional descriptive research design. The area of study was in Meru County, Imenti South Sub County. The district has been in existence since 2008 after being curved out of the old Meru Central District. It included women of 18 to 49 years in the childbearing phase attending maternal and child health clinic in level three health facilities. The researcher purposively selected the two health facilities to participate in the study since they were the key players and latest recipients of free maternity services initiative in the last five years in Imenti South Sub County. This study used both probability and non-probability sampling methods. The researcher purposively selected Kiarago and Uruku level three health facilities to participate in the study because of their location and the population they serve. The research included only women between the ages of 18 and 49 who were of reproductive age and sought treatment at Kiarago and Uruku level three facilities mother and child health clinics. The study heavily relied on data obtained primarily through self-administered questionnaires and observational checklist. A permit from National Commission for Science, Technology and Innovation was acquired as the law necessitated, and then she made a self-introductory letter to the health facilities. The data obtained utilizing the observational checklist as well as the questionnaires were evaluated by the researcher for completeness. The data was then coded with the help of SPSS program version 25. The study employed descriptive statistics analysis of data with the aid of SPSS program. Data presentation was based on bar graphs, tables, histograms, and pie charts. The approach entailed calculating the frequencies, mean, percentages, and standard deviations of data obtained to help interpret the findings.

## 4.0 Results and Findings

### 4.1 Client-related Factors and Uptake of Maternity Services

The second objective focused on examining client related factors and how they influence uptake of maternity services by women of childbearing age. In order to address the objective, the following indicators were assessed: quality of care, health care worker’s attitude, and ignorance. Statistical analysis was done and results summarized in the Table 1 below.

**Table 1:**
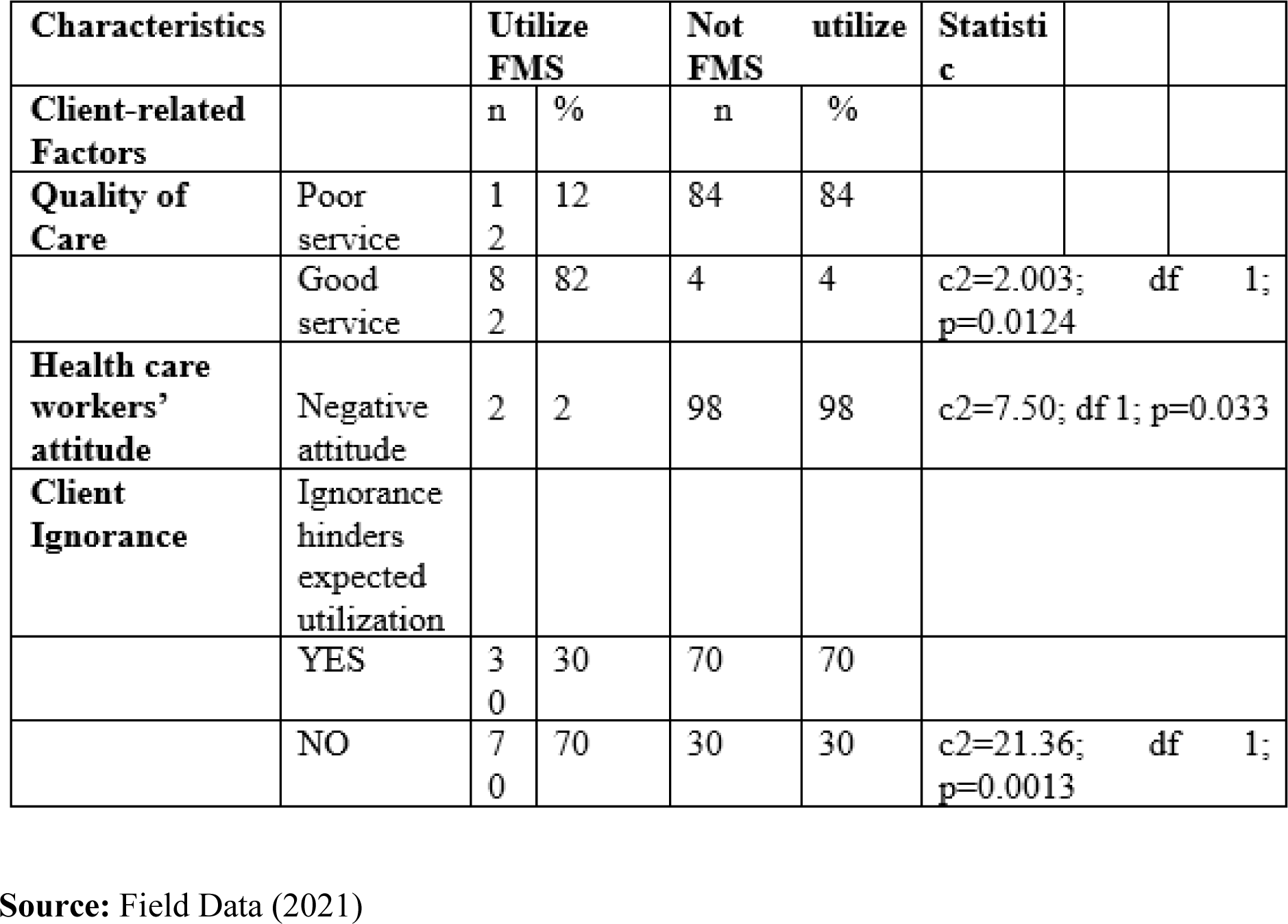
Client-related Factors and Uptake of Maternity Services.

From Table 1 presented above, 84% did not utilize free maternity services because of poor services offered. Only 12% utilized the services. From the same table, 82% said they would utilize the services if the quality of care was good and 4% said they would not utilize the services even if the quality of care was good. The table also showed how health care workers attitude affected uptake of maternity services with 98% of the respondent saying they avoided the services because of negative attitude of the health workers. 2% said they would utilize the services even with negative attitude of the health workers.

Results in Table 1 show analysis of the correlation matrix with strong positive correlations anchored by (c^2^=2.003; df 1; p=0.0124) between quality of services offered and uptake of maternity services. Further analysis was based on identifying health care workers’ attitude and uptake of maternity services. The results shows analysis of the correlation matrix with strong positive correlations (c^2^=7.50; df 1; p=0.033) between negative attitude of health care workers and uptake of maternity services. The results in Tale 4 indicate client ignorance contributes low uptake of maternity services with a strong positive correlation (c^2^=21.36; df 1; p=0.0013) between client ignorance and uptake of maternity services.

### 4.2 Client Ignorance and uptake of Maternity Services

Further investigation was made to ascertain the relationship between client ignorance and uptake of Maternity Services.

The results are reported in the following Figure

Figure 1 shows that 41.4% totally agreed, 30.3% agreed, 20.2% disagreed and 4% totally disagreed that client ignorance affected uptake of maternity services. 4% of the respondents were not aware how client ignorance affected uptake of maternity services. The results in Figure 1 indicate client ignorance contributes to low uptake of Maternity Services. Patients and providers are likely to encounter authoritarian and frightening responses particularly during childbirth.

**Figure 1:**
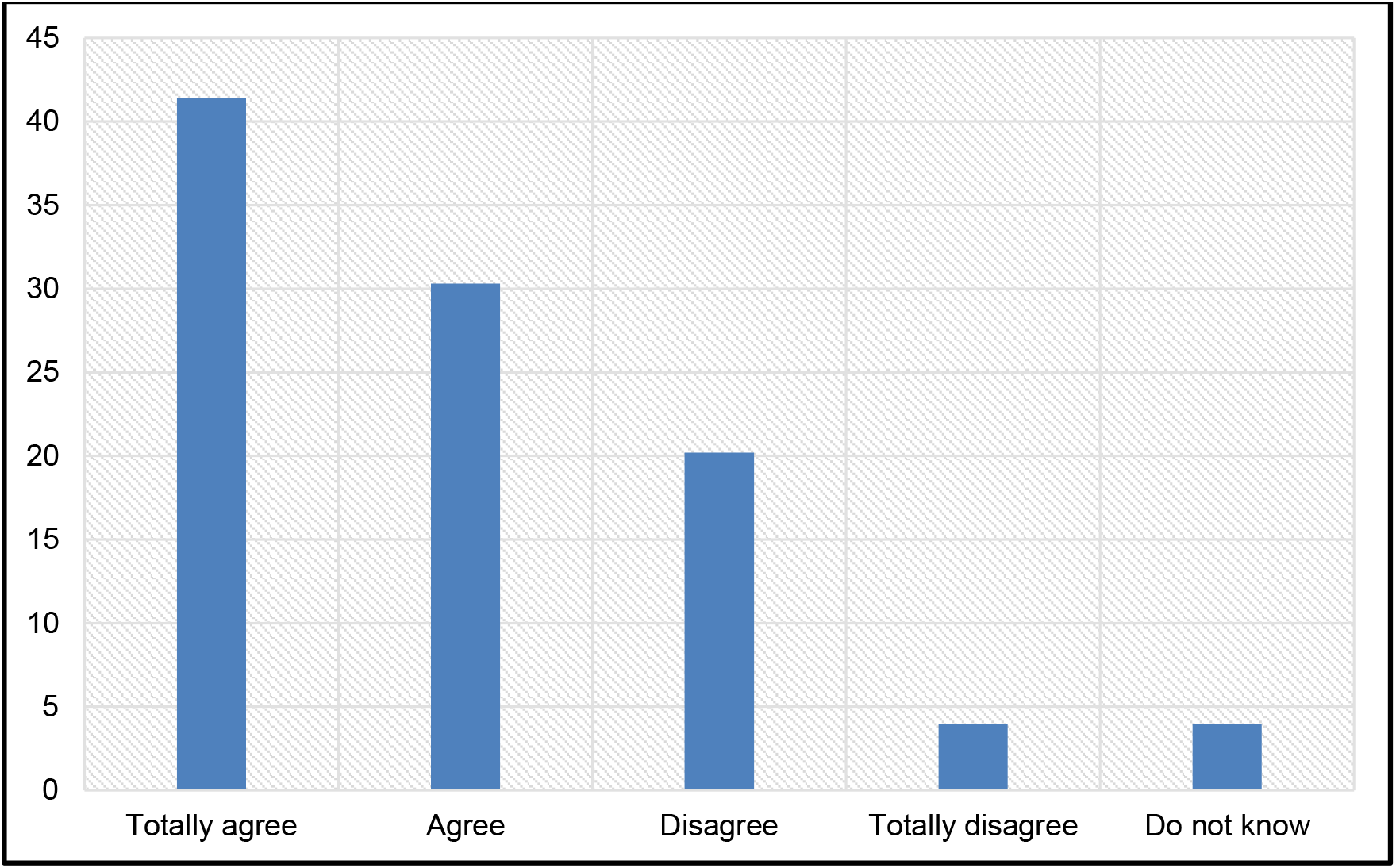
Client Ignorance and uptake of Maternity Services.

## 5.0 SUMMARY, CONCLUSIONS AND RECOMMENDATIONS

### 5.1 Summary Client-related factors influencing uptake of maternity services

The client related factors under investigation (quality of care, health care worker’s attitude and ignorance) influenced uptake free maternity services positively or negatively. The study findings indicated that quality of care offered in health facilities and attitude of the health care workers influenced women of childbearing age decision on uptake of maternity services. The findings showed that more than half of women did not utilize the free maternity services because of poor quality of services and negative attitude of the health care workers at 84% and 98% respectively. Also 44.4% of women strongly agreed that client ignorance influenced uptake of maternity services. This concurs with a study by Gathigah, (2013) that found out that even with free maternity services, uptake of government health facilities remains low. He noted that this was due to factors such as overcrowding, poor quality of care, negative health worker’s attitude and mistreatments, cultural influence and ignorance.

### 5.2 Conclusion

Free Maternal Service uptake among women of child bearing age varies with age, educational level and household monthly income. With regards to client related factors, quality of care and health care workers’ attitudes were found to be significantly associated with low Free Maternal Service seeking behavior among women of child-bearing age most of whom reported seeking Maternal Services in private hospitals. The prevalence of Free Maternity Services uptake was linked to the efficiency of health facility related-factors (facility location and health infrastructure) which influenced perceptions among child-bearing women and their uptake of Maternal Services in Imenti South Sub County level three health facilities. Women of child-bearing age tend to avoid utilizing Free Maternal Services due to their preference for private hospitals which offer better and quality services, lack of adequate facilities and negative attitude of health care service providers.

### 5.3 Recommendations

First, the government empowers the local women through access to quality education and support for rural employment opportunities. This will ensure women have alternative livelihoods with incomes and also remain informed on free maternity services access and uptake.

Second, the government should increase the effectiveness of customer satisfaction initiatives. It is important for such campaigns to take into consideration the many elements that impact attitudes and behaviors. It is possible that some of these efforts will involve a review of human resource planning, public relations, and human resource policies that will guarantee that the best service is provided.

## Data Availability

None

